# Experience of Adults with Upper-limb Difference and their Views on Sensory Feedback for Prostheses: A Mixed Methods Study

**DOI:** 10.1101/2022.03.13.22272179

**Authors:** Leen Jabban, Benjamin W. Metcalfe, Jonathan Raines, Dingguo Zhang, Ben Ainsworth

## Abstract

**Background:** Upper-limb prostheses are regularly abandoned, in part due to the mismatch between user needs and prostheses performance. Sensory feedback is among several technological advances that have been proposed to reduce device abandonment rates. While it has already been introduced in some high-end commercial prostheses, limited data is available about user expectations in relation to sensory feedback. The aim of this study is thus to use a mixed methods approach to provide a detailed insight of users’ perceptions and expectations of sensory feedback technology, to ensure the addition of sensory feedback is as acceptable, engaging and ultimately as useful as possible for users and, in turn, reduce the reliance on compensatory movements that lead to overuse syndrome.

**Methods:** The study involved an online survey (N=37) and video call interviews (N=15) where adults with upper-limb differences were asked about their experience with limb difference and prosthesis use (if applicable) and their expectations about sensory feedback to prostheses. The survey data were analysed quantitatively and descriptively to establish the range of sensory feedback needs and their variations across the different demographics. Reflexive thematic analysis was performed on the interview data, and data triangulation was used to understand key behavioural issues to generate actionable guiding principles for the development of sensory feedback systems.

**Results:** The survey provided a list of practical examples and suggestions that did not vary with the different causes of limb difference or prosthesis use. The interviews showed that although sensory feedback is a desired feature, it must prove to have more benefits than drawbacks. The key benefit mentioned by participants was increasing trust, which requires a highly reliable system that provides input from several areas of the hand rather than just the fingertips. The feedback system should also complement existing implicit feedback sources without causing confusion or discomfort. Further, the effect sensory feedback has on the users’ psychological wellbeing was highlighted as an important consideration that varies between individuals and should therefore be discussed. The results obtained were used to develop guiding principles for the design and implementation of sensory feedback systems.

**Conclusions:** This study provides a mixed-methods research on the sensory feedback needs of adults with upper-limb differences, enabling a deeper understanding of their expectations and worries. Guiding principles were developed based on the results of a survey and interviews to inform the development and assessment of sensory feedback for upper-limb prostheses.

## Introduction

Upper-limb prostheses are used to aid with activities of daily living (ADL) and improve symmetry for individuals with upper-limb differences. However, they are regularly abandoned due to the mismatch between user needs and device performance [1, 2, 3]. Users tend to find prostheses uncomfortable and difficult to use, issues that have persisted since they were reported in the 1996 Atkins survey [4]. In fact, despite significant technological advancement, there has been no significant reduction in the rate of prostheses abandonment in the last decade [5]. Individuals with limb differences who do not use prostheses rely on alternative ways to carry out ADL that do not require both hands. However, those methods tend to utilise compensatory movements [6] that, over time, can result in pain or overuse syndrome [7, 8] that can lead to reduced physical function and quality of life [6, 9].

### Sensory Feedback User Needs

The term *Sensory feedback* is used to describe sensations delivered to prostheses users to relay information about the prosthesis’ state (e.g., current grip force). Sensory feedback has been shown to be advantageous in terms of improving control, increasing embodiment and reducing phantom limb pain^1^ [12]. Despite the expanding literature on sensory feedback and its introduction to some high-end commercial prostheses, limited data is available about what user expectations [13].

Surveys related to upper-limb prostheses typically include questions investigating the interest in sensory feedback, with variable results [14, 4, 15, 16]. Lewis et al. designed a survey that *specifically* investigated sensory feedback. The results showed that 94% of respondents rely on some sort of implicit feedback (such as vision) and 88% ranked sensory feedback as important. Grip force was found to be the most important feedback, followed by movement and position. In terms of sensory feedback modalities, temperature ranked highest, followed by vibration and electric stimulation [17].

While such surveys are useful, and enable the opinion of larger numbers of participants to be captured, they do not provide the richness of data available through qualitative methods such as interviews. Graczyk et al. interviewed two participants about their experience after a home trial of implanted sensory feedback. Despite the interview including general information related to how sensory feedback could enable return to normalcy, a large focus was placed on the specific feedback system (e.g., system operation and sensation stability) [18], with limited generalisability of results. Thus, there is need for a dedicated qualitative study to investigate user needs and expectations of sensory feedback systems *before* they test one. This ensures that the user is not limited by the constraints of an already-developed system, and that the in-depth understanding user needs remain at the heart of the process throughout the stages of intervention planning, development, optimisation, and evaluation.

## Research Aim

The aim of the study was to understand the experience of individuals with upper-limb differences, their perception of the need for sensory feedback in prostheses and their expectations of such systems. This understanding will build on the available literature and will be used to guide the design of sensory feedback systems and the outcome measures used to assess those systems.

## Methods

### Study Design

A pragmatist epistemological framework was followed in which the focus of the research was to produce findings that can be effectively applied in a real world context, acknowledging the existence of both objective and subjective elements to experiences [19, 20]. A mixed-methods approach was used involving an online survey and interviews. The online survey was used to enable the views of a larger sample to be captured and compared across included demographics. Richer data was obtained from a subset of the survey participants through video call interviews, enabling the deeper understanding of individual experiences.

The survey was shared on social media and was sent to relevant charities (listed in the Acknowledgements) for dissemination. Participants who completed the survey were given the option to register interest in further participation in the study via an interview. Both the survey and interviews took place in parallel, and the survey responses were monitored to guide the interviews; however, the survey responses were anonymous and, therefore, no follow up questions were asked to the participants based on their survey responses.

This study was approved by the University of Bath’s Psychology Research Ethics Committee (20-237), and is part of a larger study using the Person-Based Approach (PBA) to guide the design of sensory feedback systems for upper-limb prostheses. The PBA emphasizes using iterative qualitative research and evidence synthesis to develop a deep understanding of key behaviours of potential users, to understand how they engage with any targeted behaviours within interventions [21]. It provides a systematic method to apply this gained understanding, by developing guiding principles and key intervention features to optimise intervention benefits. Those guiding principles are then continually updated throughout the planning, design, development and evaluation stages. The PBA values autonomy and empathetic understanding, which are particularly important for personal devices, such as prostheses [22].

### Participants

Participants were a convenience sample of adults with upper-limb differences. The sample provided a range of different causes and levels of limb difference as well as different patterns of prostheses use (or non-use). The survey recorded 37 responses (see Figure 1 for a summary of participant demographics), of which 22 registered an interest in further participation, and were invited for an interview (of which 16 accepted). Only data from 15 interviews were used due to a recording error in one. The full demographic profile of the participants in the survey and interviews can be found in Appendices A and B.

**Figure 1:**
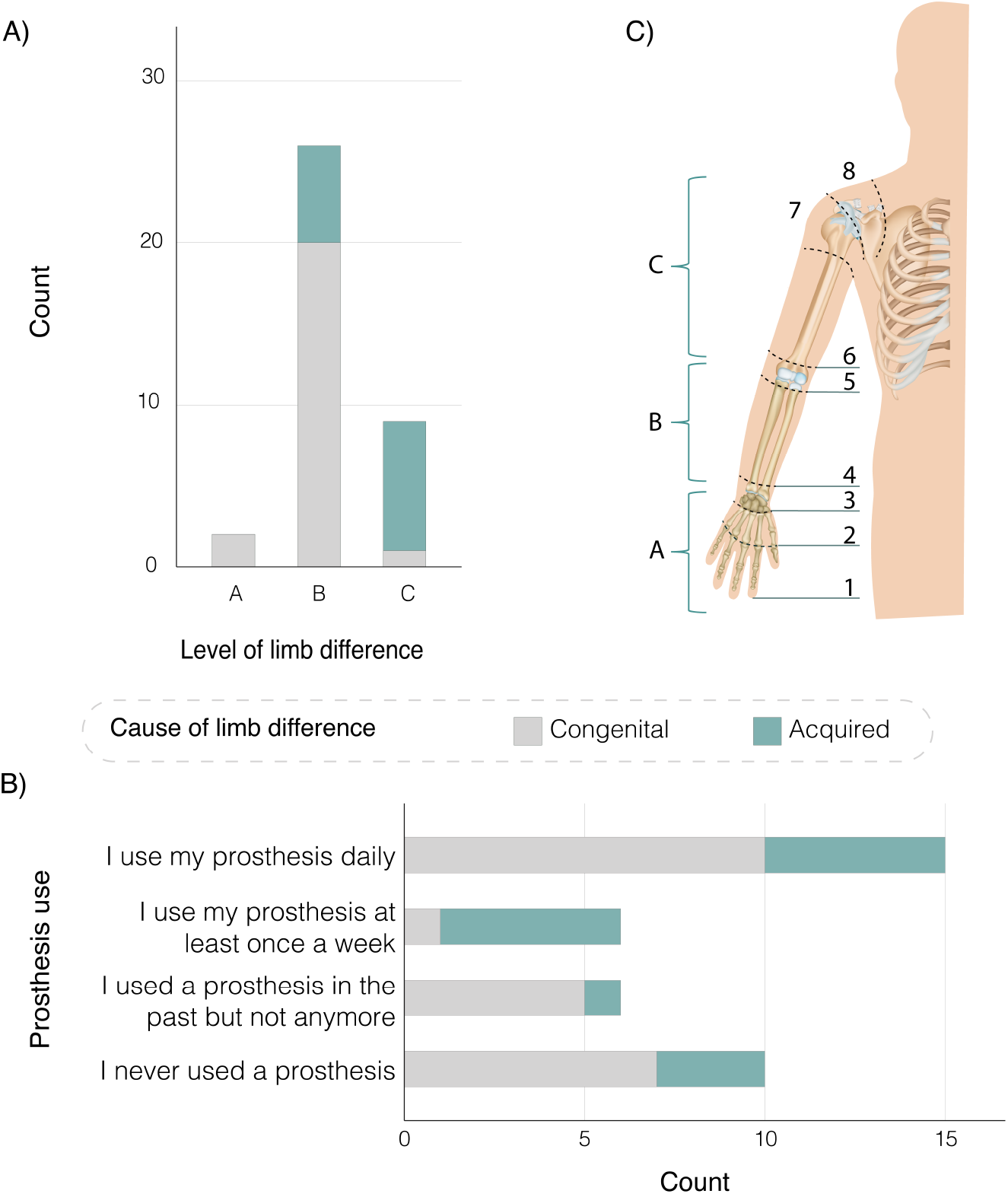
Summary of participant demographics. A)level of limb difference split based on cause. B) Types of prostheses use split based on cause of limb difference. C)Definition of the different levels of limb difference. Note that participants were shown this figure and asked to select the level that most represents their limb difference using the numbers 1 to 8. The different levels were then categorised into A, B and C.

### Procedure

#### Survey

The survey was hosted online on Online Surveys (Jisc) and was open from the 11^th^ of January 2021 untill the 31^st^ of July 2021. The survey was predominantly qualitative, by relying on open text responses, and included questions on the experience of the participants with limb difference and prosthesis use (if applicable) and their opinion on sensory feedback. Sensory feedback was described as follows:

> *We are working on developing a sensory feedback system for upper-limb prostheses. The system will rely on placing electrodes on the residual limb. Those electrodes will provide you with a sensation that you can use as an indication of how the prosthesis is engaging with the object held (for example, how hard you are holding an object)*.

A flowchart of the survey questions is provided in Appendix D. The full survey and a spreadsheet of the open text responses are available in Additional files 3 and 4, respectively.

The first draft of the survey was produced by LJ who has an engineering background. The structure and questions were then modified based on input from BA (Psychology), BWM (Biomedical Engineering) and JR (Engineering with industrial experience working on upper-limb prostheses) to ensure that the questions were relevant, easy to understand and non-leading. The survey was also piloted with two adults with upper-limb difference^2^ who were asked to complete it online during a video call while explaining their interpretation of what each of the questions means (concurrent think aloud test) [23]. An overview of the changes made can be found in Additional file 1.

### Interviews

The interviews were held online using Microsoft Teams (with LJ as the interviewer) and recording started after welcoming the participants and confirming their consent. The recordings lasted between 18 and 56 minutes with an average of 33 minutes. A semi-structured interviewing style was followed, with the list of questions shown in Additional file 2 being used as a guide. The full transcripts of the interviews are considered confidential (particularly given the small population size) and will, in line with ethical processes, not be shared beyond the quotation extracts provided.

### Data Analysis

#### Survey data analysis

The statistical analysis of the quantitative results was carried out using SPSS 27 (IBM, Armonk, NY). Open text responses were summarised into tables to represent the range of responses obtained for each question (see Additional file 1). The Kruskal-Wallis H test was used to compare the means between different groups, and the Wilcoxon signed-rank test was used to compare changes in scores. Spearman’s Correlation test was used to test the strength of the relationship between variables with a coefficient of ± 0.1 required to show a “small” correlation. A p-value of 0.05 was used as the significance level [24]. Given the exploratory nature of the study, significance levels were not corrected for multiple comparisons.

### Interview data analysis

Reflexive thematic analysis was used to generate the themes from the interview transcripts, following Braun and Clarke [25, 26, 27]. The analysis started with data familiarization during which LJ produced verbatim transcripts of the interviews using Express Scribe Pro (V 9.22, NCH Software) and noted down preliminary thoughts. Codes were generated and collapsed into manageable numbers using NVivo 12 (QSR International, 2020), and then grouped into themes relevant to the research questions. Upon reviewing the themes, the coding step was revisited (manually) before defining and naming the final themes (and sub-themes). The codes and themes were discussed with BA at the different stages and the final themes were reviewed by all authors.

### Data Triangulation

The data from the survey and interviews were analysed separately then combined to highlight the key behavioural issues and achieve the research aim to develop the guiding principles for sensory feedback systems [28]. The thematic analysis findings were used as a starting point and were cross-referenced with the survey findings, where significant overlap was found in the core issues and needs. However, the themes generated from the interviews covered a wider range of important issues; particularly showing the role that sensory feedback can have on user’s emotional wellbeing.

### Stakeholder Consultation

The final guiding principles were the product of an iterative development process during which a mix of academic, clinical and lived experience stakeholders were consulted^3^. The consultation was performed via individual video calls during which the results and preliminary guiding principles were shared and discussed. The final version is based on the comments received from all stakeholders.

## Results

This section will present the interview findings and integrate survey results as suitable to avoid repetition. Participants often mentioned that completing the survey beforehand enabled them to have some time to think about sensory feedback and how it would fit in their lives. This resulted in better examples, clearer explanations of how they envision sensory feedback to be useful and interesting discussions overall. Please note that the names used are pseudonyms.

> *“The big question is, would I find it useful? The answer is I didn’t think so until I really started thinking about it because I have never had it*.*”* (Jerome)

The interview findings were summarised into four themes (Figure 2) reflecting the key issues related to sensory feedback. All participants shared a similar approach to considering sensory feedback which involved a cost-benefit analysis *(Overarching theme: Is is worth it?)*. They were mostly worried about sensory feedback being uncomfortable or intrusive. However, they found the prospect of being able to trust the prosthesis to be a key benefit of sensory feedback that would only be possible if the system itself was highly reliable *(Will I be able to trust the prosthesis?)*. Further, participants listed examples of activities that they would expect sensory feedback to be useful with *(Will I be able to do more?)*. Many participants also highlighted how sensory feedback, and prostheses in general, can affect how they feel both about themselves and during interactions with others, which was another aspect they considered as part of the cost-benefit analysis *(How will it affect how I feel?)*.

**Figure 2:**
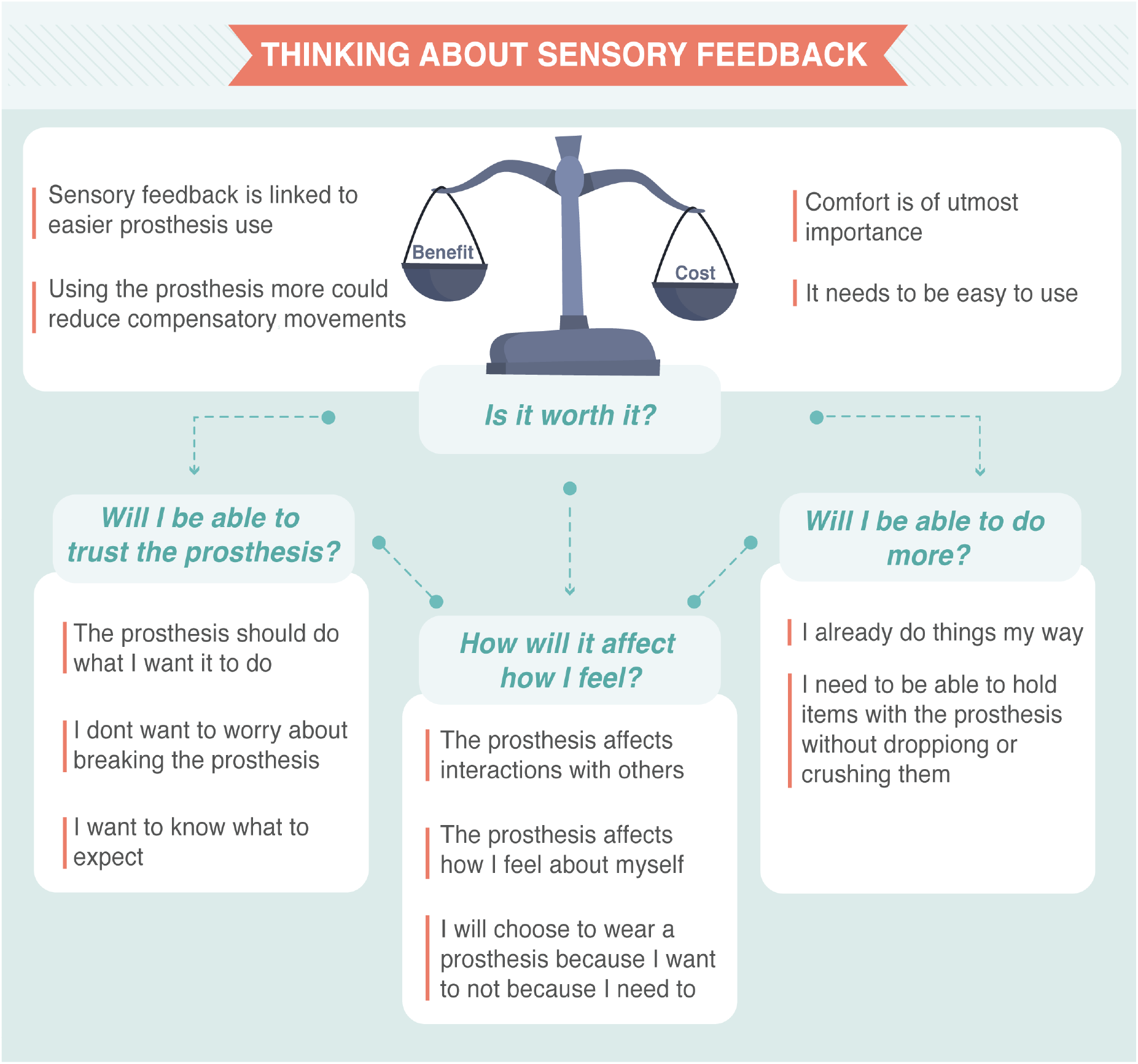
Map of the themes generated from the interviews with adults with limb differences on their experience and what they expect of a sensory feedback system. The themes are presented as the main questions participants considered when thinking about sensory feedback, and prostheses in general. Sub-themes are presented to further explain the themes.

The following subsections will elaborate on the highlighted themes and subthemes and include excerpts from the interviews, with […] used to mark omissions and ** to indicated words that the participants emphasised. Relevant survey findings that support the different themes are presented throughout, and the full survey results can be found in Additional file 4.

### Theme 1: Will I be able to trust the prosthesis?

Trust, in the context of the user not being able to trust that the prosthesis is doing what they expected it to, was mentioned repeatedly. This lack of trust made prosthesis use limited and unenjoyable. Survey responses provided specific examples, where users reported worrying about safety when holding babies and shaking hands. The most common reported error (37.5% of survey participants) was related to holding objects securely without dropping or crushing them. Feedback was thought to increase trust by enabling the user to know what the prosthesis is doing.

> *“just trusting the device*.[… *] if a fully-abled person was using one of those gripper things to lift certain objects, it would be a bit nervous or you would be a bit nervous, but if you could eliminate that nervousness and it was just natural, it would be*… *It would just be more kind of fluid and fun and just kind of enjoyable to sort of do everyday tasks*.*”* (Tony)

#### A) The prosthesis should do what I want it to do

The uncertainty associated with prosthesis use, and myoelectric control in particular, made participants feel like they have no control. This lack of control and trust lead to reliance on vision, which the participants wanted to reduce. Any new feedback modalities must be considered trust worthy, for example in terms of both consistency and latency. Participants frequently expressed valuing reliability over an increased number of functions.

> *“one of the other reasons I don’t wear [the prosthesis] all the time [*…*] say it’s like 90 or 95% reliable but then it just like fails 5% of the time*.[…*] I always like have to second guess like, oh, am I going to be able to use this in the way I expect?[*…*] how reliable are sensors, do they cut out? If I’m using this to be able to see like, oh, like I’m confident that I’m not going to drop something. Um, then it, I think it just has to have very high reliability for me to have confidence in it*.*”* (Kevin)

#### B) I don’t want to worry about breaking the prosthesis

In addition to functionality, prostheses fragility can lead to trust issues associated with the fear of damage, which is exacerbated by their high cost. The addition of sensory feedback lead some participants to worry about increased device complexity, such as new pressure sensors, which could lead to more failures.

> *“well that is another thing that can go wrong, or it is another thing that won’t work or it is another thing that will wear out*.*”* (Guy)

On the other hand, sensory feedback can help diminish the concerns associated with device damage by enabling rapid assessment of the applied forces.

> *“That is another reason why I want to get feedback for it*.[…*] if I felt some sort of feedback like oh you’re hitting your fingers I would be like oh I better stop that because I’m not trying to break it*.*”* (Ted)

#### C) I want to know what to expect

Several participants demonstrated interest and curiosity in experiencing sensory feedback, and new prostheses in general. This was often due to the perceived futuristic nature of prostheses and the participants’ view of them as “gadgets”. However, some found uncertain expectations to be worrying.

> *“What to expect putting it on? I think I would have been quite nervous [laughs] like what’s gonna happen when you hold something*… *“* (Grace)

Describing the functionality of a prosthesis builds expectations that should be managed carefully. Participants who had high expectations of what the prosthesis can offer, especially given its cost, were disappointed. This highlights the importance of clear, effective and coherent communication with the rehabilitation team to understand what the user wants and expects.

> *“when I got my other prosthetic arm it was like just a absolute, like kick in the face, because I was like, this is useless. Like, this is not what I wanted it for*..*and they weren’t really listening to me in the centre, either*.*”* (Laura)

Expectation management is particularly relevant when describing sensory feedback. The user should be made aware of the limitations; both in terms of quality of sensation and the locations of the sensors. Unattainable expectations were evident from the interviews, with several participants expecting to feel different textures from all over the prosthesis, despite the sensations being described in the survey as *“similar to the feeling of your phone vibrating when you get a notification”*. It was suggested that the term *“sensory feedback”* might contribute to those high expectations and should be replaced with *“tactile feedback”* when vibrotactile and electrotactile feedback methods are employed.

> *“I think it would be amazing to see how it feels with the sensations. [*…*] whether it be touching a table like you get that feeling with that or maybe petting an animal you know that kind of thing*.*”* (Kelsey)

Similarly, being upfront about the importance of training is important. While most participants acknowledged that time would be needed to learn how to use the system, they frequently expressed interest in specific training to aid with, and accelerate, the process. This was often guided by their experience with prosthesis use becoming easier with time, as evidenced in the survey where participants reported a significant reduction in the mental effort required to use the prosthesis after practice (Mean change = −1.38, Z=-2.418, p=0.016, see Figure 3). However, no correlation (Spearman’s Correlation Coefficient= −0.042, P=0.860) was found between the duration of use and the reduction in *perceived* mental effort. It is possible that participants who used their prosthesis for a long time have a reduced perception of how difficult it was to use when they first got it due to recall bias (Spearman’s Correlation Coefficient =-0.324, p=0.164), which is influenced by the recall period and can result in underestimated association [29].

**Figure 3:**
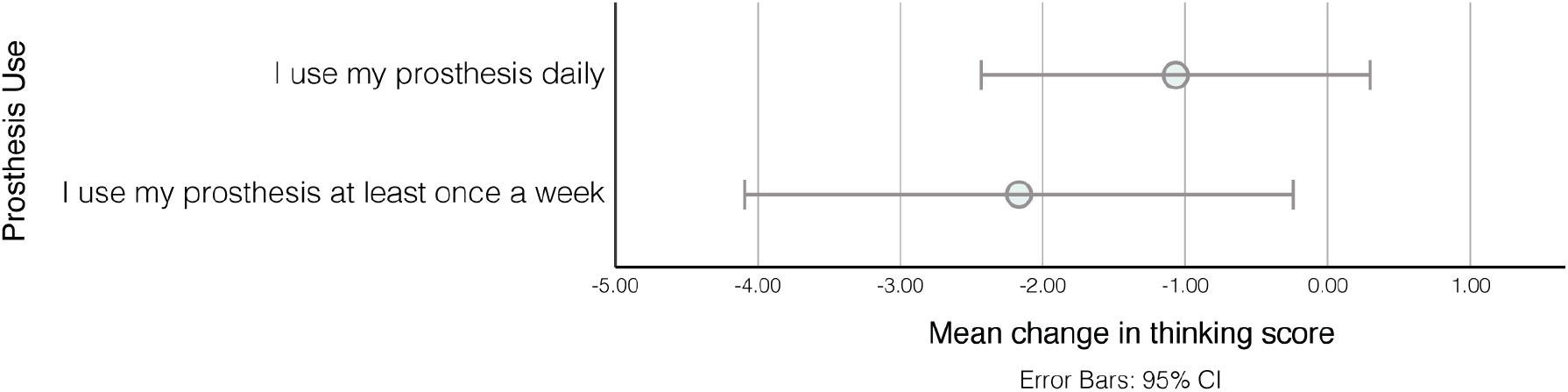
Comparison of the mean change in the thinking score between participants who use their prostheses daily and those who use it once a week, showing the 95% confidence intervals. The scoring system was a scale between 0 and 5 with 0 indicating *“I can use the prosthesis and maintain a conversation without interruption”* and 5 indicating *I cannot do anything else while trying to use the prosthesis”*.

> *“I think it might take a while for someone to get used to [the feedback]. I guess you would need some sort of activities to. Well, I like a learning path for them to work through*.*”* (Tony)

### Theme 2: Will I be able to do more?

As well as increasing trust, all participants expressed wanting something *“functional”* that increases the number of tasks they can carry out or makes the tasks easier. The tasks were mostly bi-manual activities (wherein the prosthesis is used to hold an object being manipulated with the other hand) or general holding of items. Prostheses were used when they *increase* functionality. It is expected that a similar trend will be observed with sensory feedback where, for it to be used effectively, users should be able to see how it *improves* what they can currently do.

> *“sometimes when people go to design, they think that*… *they forget that the person will have worked around and find their own way of doing certain things and I think it’s very important*.*”* (Tony)

#### A) I already do things my way

Participants explained how they relied on planning ahead to do things their way, depending on their level of limb difference and day-to-day needs. Participants with congenital limb difference highlighted that, being born that way, they did not have to adapt to different ways of carrying out ADL.

> *“When I was learning to crawl. I didn’t know there is another way to crawl, I just figured out how to crawl and same thing with like everything else. When you learn the mechanism to make your body work*… *I never had a new challenge to learning it, I just learned it*.*”* (Ross)

The reliance on alternative ways of carrying out ADL lead some participants to reject prostheses because they did not need it, and others to abandon it because it got in the way, resulting in lost functionality, rather than gained.

> *“for me, it was almost wearing a third, a third appendage or a third limb that kind of bumped into things*.*”* (Georgie)

Understanding that users have developed their own ways to carry out tasks is directly relevant to sensory feedback. The survey showed that, despite not having any explicit feedback, 74% of those who tried a prosthesis (and 86% of current users) actively rely on looking at the prosthesis, feeling the vibration through the socket or listening to the sound of the motors (Figure 4). Therefore, sensory feedback should be designed to *complement* those sources, rather than replace them.

**Figure 4:**
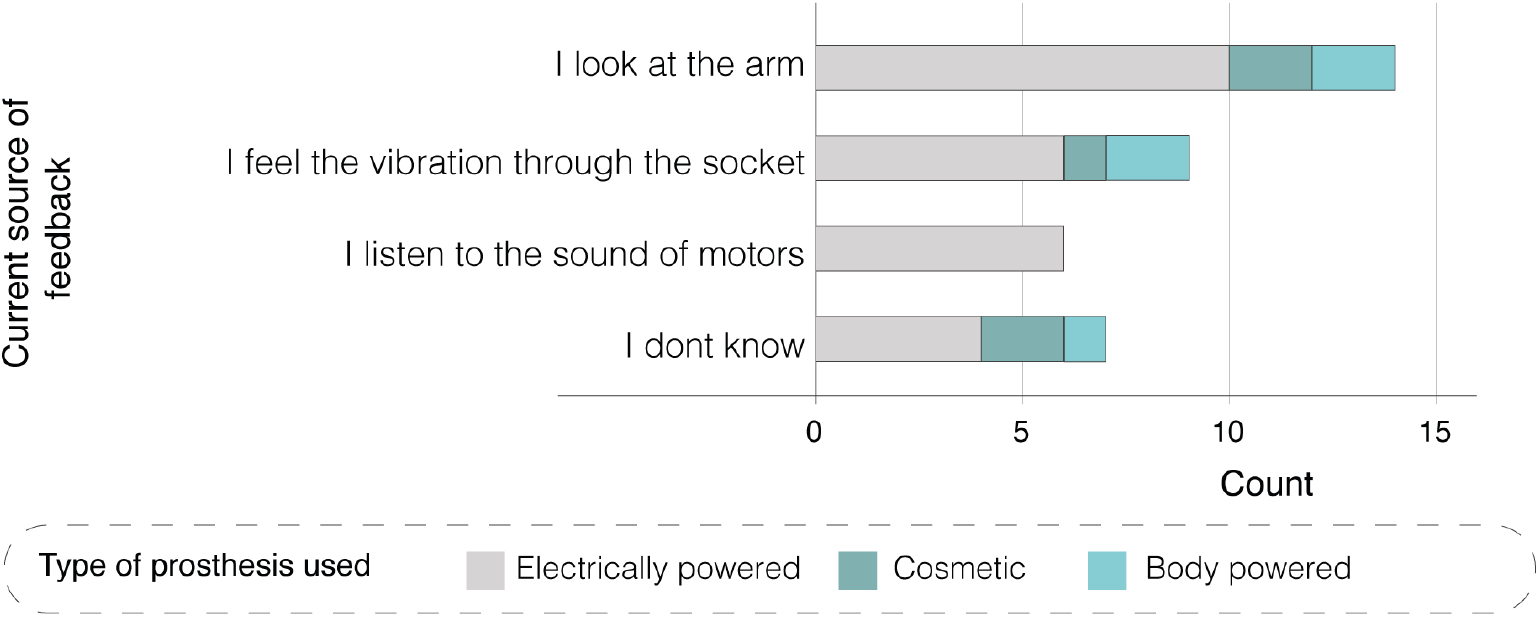
Sources of implicit feedback showing the split based on the type of prosthesis used.

#### B) I would like to be able to hold items with the prosthesis without dropping or crushing them

The most frequent task associated with prosthesis use was picking up and holding objects when the other hand is busy or when the object needs to be held away from the body. This can be difficult to do with fragile objects due to poor grip force control without feedback. Pressure feedback was thought to make this easier if it was reliable, repeatable and placed at suitable locations on the hand. Those locations should not be limited to the fingertips (as not all grips result in the fingertips touching the object).

> *“there are things that I hold that touch my palm more than my fingers, especially depending on what shape it is because you’re not always going to hold something with your fingers”* (Jessica)

### Theme 3: How will it affect how I feel?

Whilst functionality dominated the discussion, the psychological effect of wearing a prosthesis was highlighted as a key consideration. The effect the prostheses have on psychological well-being was more evident during the interviews than the survey where participants rated emotional benefit as the lowest priority compared to improving control, increasing embodiment and reducing phantom limb pain (see Figure 5). It should be noted that the interpretation used to describe embodiment in the survey (Makes the prosthesis feel more like a part of the body rather than a tool) is one of several used in the literature. The reader can refer to [30] and [31] for more details.

**Figure 5:**
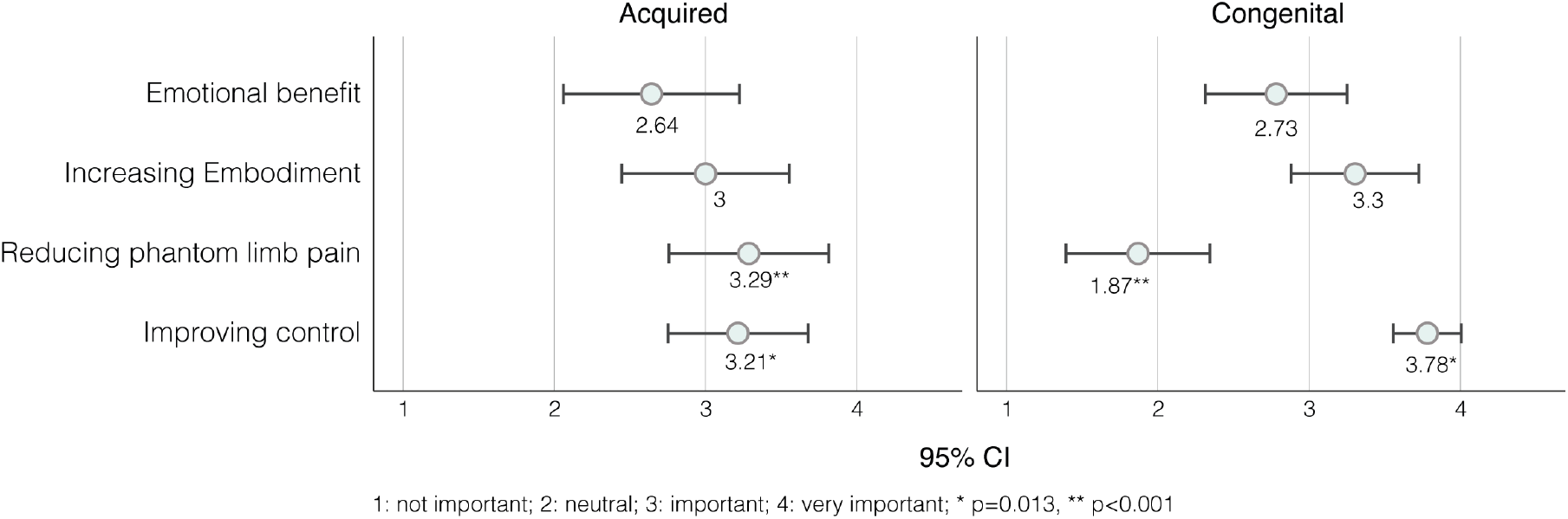
Comparison of the mean importance score of each of the sensory feedback benefits between participants with acquired and congenital limb difference, showing the 95% confidence intervals. The scoring system was a scale between 1 and 4 with 1 indicating *“Not important”* and 4 indicating *Very important”*. The p value markers compare the importance scores between participants with acquired and congenital limb difference. The description of some benefits was simplified as follows: Emotional benefit was described as: “It is emotionally pleasing to feel touch through the prosthesis”. Embodiment was described as: “Makes the prosthesis feel more like a part of the body rather than a tool”. Improving control was described as: “Makes it easier to control the prosthesis”.

> *“before you start engaging in activity, you know*.. *its being comfortable with yourself and being comfortable with the way people perceive you”* (Walter)

#### A) The prosthesis affects interactions with others

Participants talked about generally positive interactions with others, and friends and family in particular. However, several pointed out that kids’ honesty can sometimes be hurtful, and that the prostheses enabled a more positive interactions with kids due to their “coolness factor”. Prostheses had a similar effect on interactions with adults as well, facilitating the conversations on limb difference and shifting the focus onto the device itself. A few mentioned that they found it easier to be accepted and included when wearing a prosthesis. This was mentioned in the context of both passive and myoelectric prostheses.

> *“I think that’s a great thing about these devices that it, it sort of makes it a more interesting way to have a conversation about it, I guess”* (Tony)

However, some mentioned that those conversations can get repetitive and they would sometimes prefer not to draw attention to themselves. The opinion on this matter varied between participants with some being motivated to spread awareness about limb difference and others wanting to be seen for who they are, aside from being limb different. Despite those differences, most participants mentioned occasions where they hid their limb difference or prosthesis to blend in. This highlights the importance of feedback being discrete so as not to increase the curiosity of people and lead to further questioning.

> *“I’ve gotten to a point now where I do love my arm, I love people seeing it but I also sometimes just want to blend in. I don’t need people to be like* … *wow that is a cool arm*.. *I’m like hmm cool I’ve got stuff to do”* (Ted)
>
> *“I just think that my existence as a disabled person does need to be a learning experience for people [*… *] a lot of people don’t even like, ask me my name or how I am, they literally just started like, that’s a cool hand, like tell me about it or whatever and there’s just, there’s so much more to me than my hand and I just think it’s disrespectful*.*”* (Jessica)

Some participants who do not currently wear a prosthesis wondered how others would react to the change, and some mentioned that their family might want to try the sensory feedback technology themselves, if they are able to (if it was a wearable cuff, for example).

> *“only my parents and my siblings are the people that*.. *that have known me when I wore a prosthetic before, so it would be it would be very*… *hmm*… *It would be a little different. “* (Georgie)
>
> *“Hmm [laughs] I think that if they would be able to they would probably want to try it on themselves and feel how it feels with the different sensation feels and the gripping feels of it*.*”* (Kelsey)

Another aspect of interactions with others is touch (handshakes in particular). Feedback was thought to be useful to enable the user to shake people’s hands with confidence while maintaining eye contact and not worrying about holding too tight. Even when the prosthesis is not used to shake the other person’s hand, it is useful to have it to easily transfer objects between hands with confidence and minimal visual attention.

> *“I suppose if you like shaking hands with somebody, then if [the prosthesis provides feedback] you’d know you’d actually make contact because when you shake hands with somebody, you don’t look at their hand and make sure your hand goes into their hand. Yeah. You’re more looking at their face. So that would be very useful*.*”* (Guy)

#### B) The prosthesis affects how I feel about myself

A prosthesis was thought to improve self-reliance and confidence carrying out different tasks in public. However, when the performance did not match expectations it was seen as disabling.

> *“It didn’t feel like it assisted me in terms of like doing an activity, it always felt more disabling because of the restricted function it had*.*”* (Grace)

Further, participants highlighted the importance of aesthetics. This was rarely linked to natural-looking prostheses but rather to visually pleasing, “sleek” looking devices. Cosmetic prostheses were sometimes viewed negatively as they are “not fooling anyone”.

> *“I hate the hook so much, because they’re not, they’re not aesthetic at all, but they’re also not particularly functional. So they just look horrible. Yeah, it kind of, I never felt nice. Well, I guess it’s like clothes isn’t it? When you put them on, you’ve got to feel kind of nice. And with that you just feel like something from a medical book, I guess? I don’t know. not nice*.*”* (Grace)

Prosthesis use was also linked to acceptance, more actively so with participants with acquired limb difference as they described acceptance to be key to moving on with life after limb loss. Viewing the prosthesis as a tool, rather than a replacement, and understanding its limitations was a critical part of acceptance. During the interview, Margaret seemed to be opposed to the idea of sensory feedback due to fear of how it will affect her acceptance of her limb loss.

> *“it would be useful in some senses [*…*] but I don’t know that I really want it to be honest [*…*] I suppose it’s to do with my acceptance that I am an amputee. you know, my hands, they’re not going to grow back [*…*] it’s about getting on with life without them*.*”*

When asked to elaborate on her response, she mentioned that sensory feedback might affect the alternation between different prostheses. She used multiple prostheses for different types of activities and thought that if one of them resulted in an increased sense of embodiment, she might find using the other prostheses less enjoyable.

> *“there is no one prosthetic that can replace my hands*.[…*] So I’m always going to be changing between different tools and I think if I had feeling sensation in one of them, that could make the adjustment to not having them even harder*.*”*

It is important, therefore, to ensure that feedback is described as a way to make the prosthesis more useful, rather than as a way to regain a sense of touch. It is also advisable to discuss other prostheses and tools used to assist with ADL and how sensory feedback might affect their use.

> *“I think it’s that mental thing where you have to understand it’s a tool*.[…*] it’s never going to replace your feeling of your fingertips”* (Paul)

#### C) I will choose to wear a prosthesis because I want to not because I need to

Individuals with congenital limb differences who used prostheses as children showed some rebellious behaviour against prostheses. This was often linked to being told they *need* it, which resulted in them finding alternative ways to carry out tasks to prove that they don’t. This attitude changed in adult life when they began to view prostheses as tools for specific tasks, rather than devices to rely on.

> *“my parents essentially said, well, you can’t do these certain things without it, so you have to wear it. So I kind of taught myself how to do things with pinning my shoelaces between my elbow, tying my shoes and all of that. And then by the time I was a teenager, they kind of told me I didn’t have to wear it anymore*.*”* (Georgie)
>
> *“everyone has acknowledged that I don’t *need* it. It’s just something I’m trying out and seeing if I really like it or not right now. So yeah, a lot of acceptance, which is good*.*”* (Jessica)

### Theme 4: Is it worth it?

The overarching theme of the conversations was “Is it worth it?”. While sensory feedback has several advantages, its drawbacks need to be considered and clearly communicated. Also, in the same way that prostheses are only used when useful or needed, feedback might be switched off when it is not aiding, or even hindering a task. This thought process aligns with what has been reported previously in relation to general prosthesis use [2, 32, 33, 34].

#### A) Comfort is of utmost importance

Supporting the emphasis on the importance of comfort in the literature [35], the survey showed that comfort, along with not needing a prosthesis, were the most common reasons for not using a prosthesis (see Figure 6). Comfort was also mentioned repeatedly in the interviews as a key requirement for sensory feedback. All participants described being worried about excessive and uncomfortable feedback - described as high intensity and leading to potentially painful sensations in sensitive areas, or feedback that is present for prolonged periods. Those worries were evident in the survey responses as reasons to switch the feedback off as well as justifications for specific design features (see *Opinion on sensory feedback design* in additional file 1).

**Figure 6:**
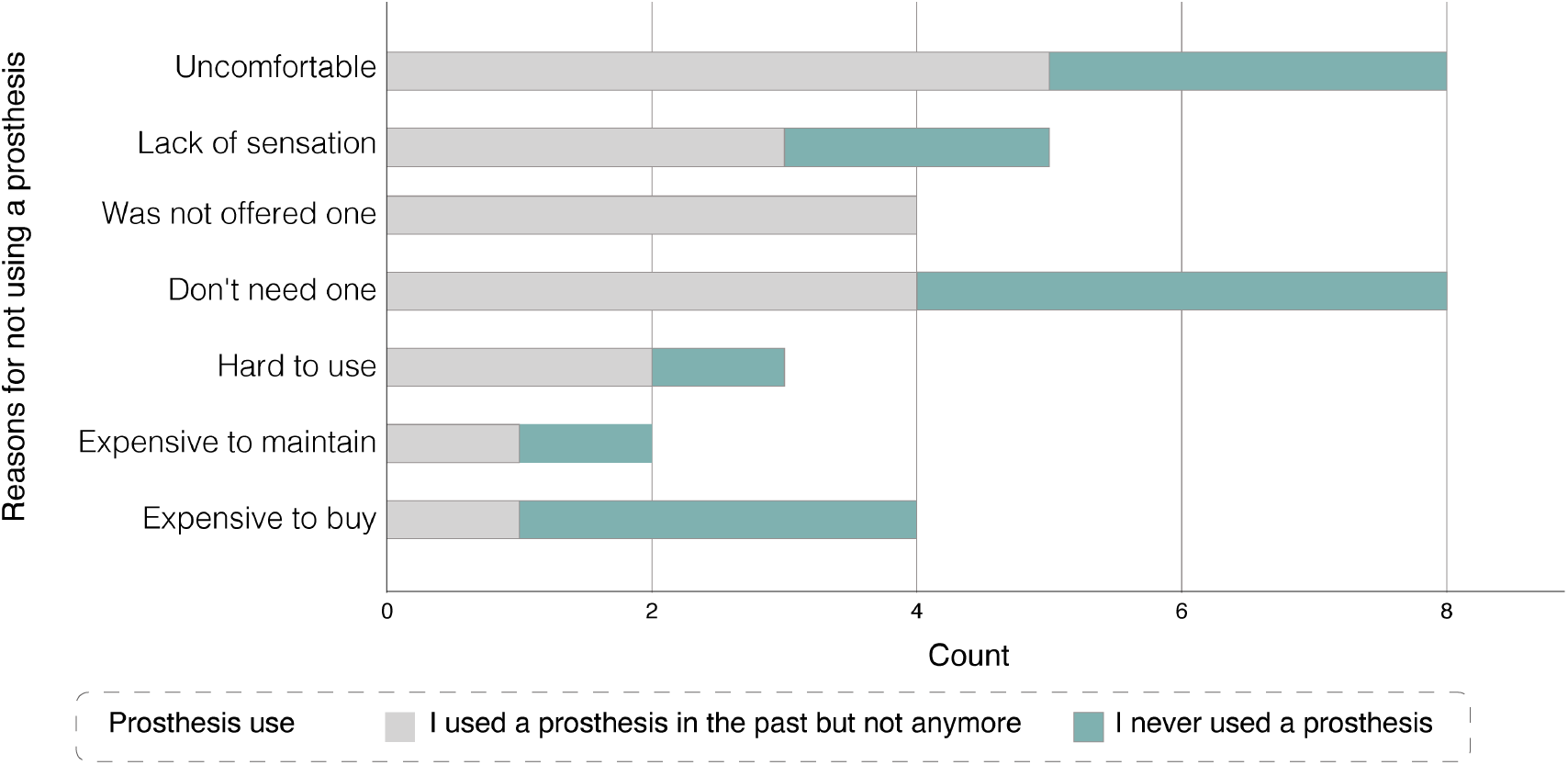
Reasons for prosthesis abandonment or non-use.

Many participants tried to come up with ways of ensuring the feedback is comfortable, e.g., reducing intensity with time. However, they often acknowledged that this is difficult and may require more advanced technological innovations. Regardless of how feedback is designed, participants expressed their interest in being able to adjust the feedback to match their comfort levels.

> *“if you’re doing the same kind of task for a while you don’t want to be told constantly-you are doing this! you’re doing this! Oh yes, I am aware! so maybe start strong and then it either weakens or just completely turns off after a while*.*”* (Tony)

Another aspect of comfort is the effect of sensory feedback on phantom limb pain. When given a list of potential sensory feedback benefits that includes: “Makes it easier to control the prostheses”, “Reduces phantom limb pain”, “makes the prosthesis feel more like a part of the body rather than a tool”, “It is emotionally pleasing to feel touch through the prosthesis”, survey respondents with acquired limb difference selected reducing phantom limb pain as the highest sensory feedback priority (see Figure 5). However, one participant mentioned that trying sensory feedback *“exacerbates [his] phantom pains beyond any measure”*. A similar comment was made in the study by Engdahl et al. where P16 reported that an experiment on sensory feedback increased their phantom limb pain [36]. It is unclear to the authors why sensory feedback may have *increased* phantom limb pain, particularly with the evidence available to the contrary [37, 38, 39, 40, 41].

#### A) It needs to be easy to use

Ease of use also influenced prosthesis usage, and in the context of feedback, intuitiveness and repeatability were the main concerns. Participants were asked their opinion on the use of a mobile application to adjust sensory feedback settings. Some participants suggested this would be a desired feature but highlighted the importance of ease to access, with Jessica suggesting that a widget^4^ option would speed up the process of adjusting settings. The reader can refer to Table 5 in additional file 1 for a summary of concerns and desired app features obtained through the survey.

> *“Like if it had an option for widgets so you didn’t even have to like log in on your phone*.*”* (Jessica)

#### C) Sensory Feedback is linked to easier prosthesis use

Participants expected that sensory feedback would make the prosthesis easier to use by creating a more natural flow of tasks, and reducing the planning and focus required. With time, repeatable feedback was expected to aid with learning, with some participants expecting not to need feedback after a few years, once they’ve mastered how to control the prosthesis.

> *“Maybe you’ve got something so sophisticated that as you do it it tells you and then you start to learn how much pressure you put on*.*”* (Jerome)

#### D) Overuse syndrome is a motivator to wear a prosthesis

Overuse syndrome was something nearly all participants mentioned or were aware of. In fact, it was one of the reasons many individuals with congenital limb differences started considering getting a prosthesis at an older age.

> *“I feel like if I could somehow balance that load, even just a little bit it might give me a longer life with my right hand and so I am *really* nervous [*…*] that’s my primary motivation-trying to figure out a way to preserve what I have and get some more functional usage*.*”* (Ross)

## Discussion

The survey and interview results demonstrated how the experience of participants influenced their needs and expectations of sensory feedback systems. Participants acknowledged the importance of reducing compensatory movements, and appreciated ways of making the prosthesis more useful to encourage increased used. They found the lack of trust in what the prosthesis is doing to be limiting, and expected reliable sensory feedback to increase this trust. However, they were wary of what adding features may do to reliability, size, and comfort. Comfort was mentioned in particular as a concern with sensory feedback, and fear of it becoming intrusive was mentioned by most participants. The effect of feedback on how the participants felt about themselves and during interactions with others was also mentioned both in terms of being a desired outcome and a potential worry.

### Implication for sensory feedback design and implementation

The findings of this study can be applied to different aspects of the design and implementation of sensory feedback systems for upper-limb prostheses by encouraging the designers and clinicians to ask the questions highlighted in Figure 7. The PBA was followed by integrating the results and existing literature to produce the *guiding principles* in Table 1. The in-depth mixed methods data allowed the identification of key user behavioural characteristics that would impact participant engagement with the intervention (sensory feedback), namely the ability to (i) trust it, (ii) see its benefit and feel empowered and (iv) comfortable using it. These characteristics were then used to define the design objectives needed to overcome the challenges linked to the behavioural characteristics. Finally, specific key design features were hypothesised to achieve the defined objectives [21]. Both the design objectives and key design features (i.e. guiding principles) can be used and modified throughout the implementation and optimisation stages of the design of sensory feedback systems and provide a valuable baseline to evaluate the intervention against to ensure that the user needs are at the heart of the process. For example, given the importance of trust on user acceptance (first design objective), designers may want to consider whether the feedback elicited is stable and repeatable during different grips, and when the arm is moved or oriented in various positions. This is particularly important in the context of myoelectric prostheses as their control can be unpredictable in some arm positions due to the change in the electrode-skin interface [42]. A feedback system that is similarly unpredictable will fail to increase trust and therefore may be rejected or not used regularly. The rest of the design objectives and features can be used in a similar manner to guide the design, assessment and implementation of sensory feedback systems.

**Table 1:** Guiding principles for the implementation of sensory feedback in upper-limb prostheses

| Design Objectives | Key Design Features |
| --- | --- |
| The sensory feedback system should enable the user to trust it | • The features of the sensory feedback system should be communicated clearly in terms of sensation quality, sensitivity and area covered<br>• Feedback sensation should not be influenced by external factors (e.g. arm position)<br>• The sensory feedback system should be durable<br>• The sensor locations should cover all of the areas used to hold objects (i.e not just the fingertips)<br>• The calibration and settings adjustment options should be designed to ensure reliability (e.g. have simple and advanced options to ensure the saved setup is not deleted by mistake) |
| Sensory feedback should be comfortable for long-term use | • Allow the user to adjust the feedback intensity and extent (i.e. how much feedback they want)<br>• The feedback should not be intrusive when objects are held for a long time<br>• The weight of the system should be negligible<br>• The user should be able to switch the feedback off easily (e.g. button on the prosthesis rather than through an app) |
| Sensory feedback should not have a negative impact on normal prosthesis use | • Sensory feedback should not distract from other activities (e.g. introduced gradually to allow the user to get accustomed to it)<br>• Sensory feedback should complement other implicit feedback sources<br>• Sensory feedback should integrate with existing notifications available through the arm (to not be confusing) |
| Sensory feedback should not draw extra attention | • Sensory feedback should not be audible<br>• The design of the sensory feedback system should either be integrated into the prosthesis or hidden without adding bulkiness (as that would cause difficulties with long-sleeve clothing) |
| The feedback should not interfere with the user's acceptance of limb difference | • The way sensory feedback is described should be appropriate to its capabilities (e.g. tactile feedback for vibrotactile feedback)<br>• The language used should be appropriate with the fact that the prosthesis is a tool and is not attempting to replace a body part (e.g. instead of asking how "natural" the sensation felt, could ask how "comfortable" it was) |

**Figure 7:**
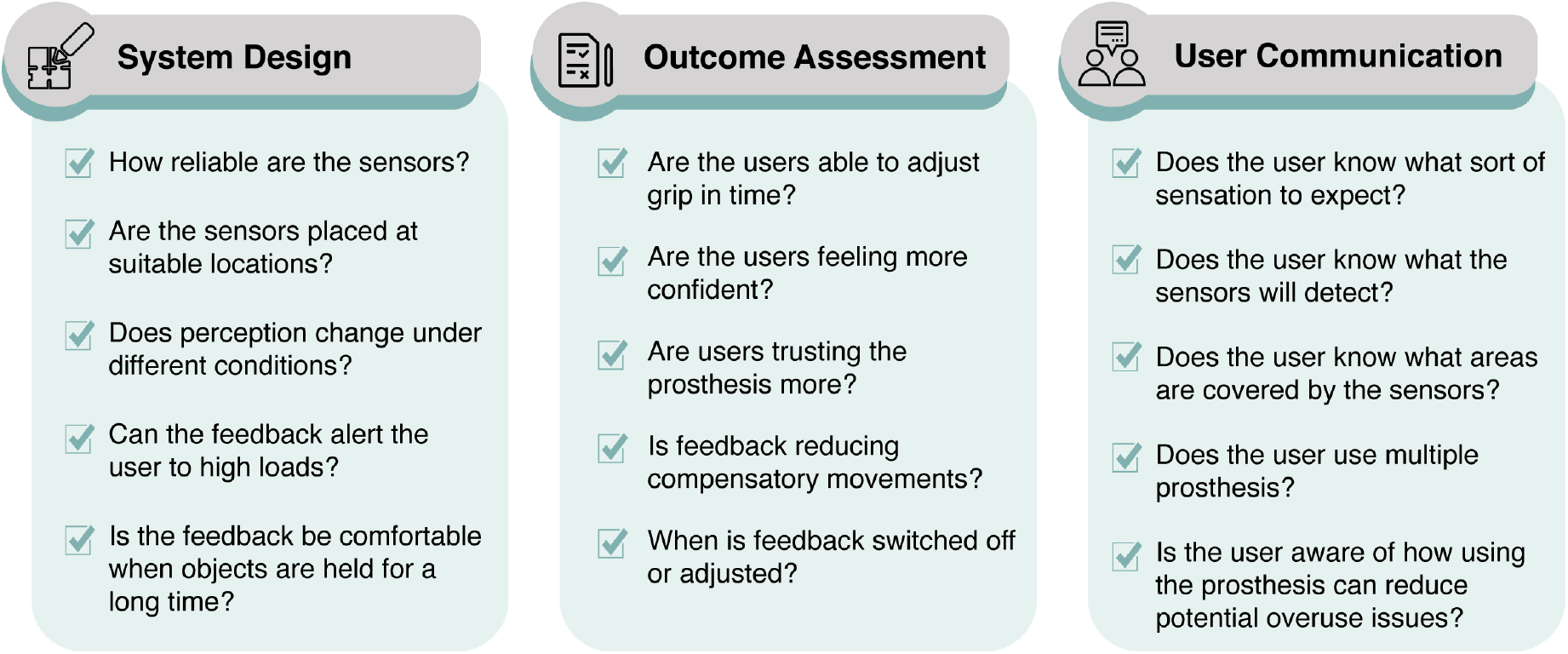
Questions that should be asked when designing, assessing or communicating to users about sensory feedback.

### Strengths and Limitations

The main strength of this study is its specific focus on sensory feedback and the use of data triangulation where both quantitative and qualitative results were combined to establish a deep understanding of the expectations of adults with upper-limb differences of such systems. The study benefited from a team with a range of experiences covering engineering, health psychology and upper-limb prosthesis (manufacturing/ clinical) experience to ensure that the recommendations accurately reflect the results whilst also being of practical use.

The study focused on vibrotactile feedback to enable the participants to conceptualise how the system would operate. The participant demographic included a high percentage of participants with congenital limb difference (Survey: 62%, interviews: 80%). However, the survey showed negligible differences in the practical experiences of participants with congenital and acquired limb difference, except for phantom limb pain (matching what was observed by Widehammer et al. [43]). The differences between acquired and congenital limb different participants were only apparent when discussing the psychological effect of adding sensory feedback, as explained in the results. Nonetheless, the developed guiding principles are expected to apply to all types of feedback and upper-limb difference causes as they focused on the core considerations rather than specific design requirements. Finally, although the study aimed to recruit participants from different demographics, the sample may not be fully representative of the population of upper-limb different adults. As with most similar studies, the participants demonstrated strong can-do attitudes [22] which may reflect a selection bias where adults that agree to participate in research are often more accepting of their abilities and are willing to engage. With the awareness of the subjectivity interview data but given the principles of information power [44], the sample size and study results were considered satisfactory.

## Conclusion

This mixed-method study highlighted the expectations and needs of adults with upper-limb differences. The survey (N=37) and interview (N=15) results are in agreement that the main perceived benefit of feedback is related to increasing the trust in the prosthesis and, thus, enabling the user to feel more confident using it to carry out ADL. The survey provided a wide range of specific practical examples to consider when designing feedback systems, while the interviews enabled a deeper understanding of the user expectations and shed light on how sensory feedback can affect how users feel about themselves and interactions with others. They also showed the different elements of the cost-benefit analysis when considering sensory feedback. The findings were translated into practical guiding principles that can be used to increase the effectiveness and impact of the developed sensory feedback systems.

## Supporting information

Additional file 1: Additional Survey information

Additional file 2:Interview schedule

Additional file 3: Survey questions

Additional file 4: Survey open text responses

Additional file 5: Interview codes

## Data Availability

All of the available data is found in the additional material. The interview transcripts are treated as confidential and will
not be shared.

### Appendix

#### A: Participant Demographics

**Table 2:** Survey and Interviews Participant Demographics. Refer to Figure 1 for the definition of the different upper-limb difference levels.

|  |  | Survey<br>(n) | N=37<br>(%) | Interviews<br>(n) | N=15<br>(%) |
| --- | --- | --- | --- | --- | --- |
| Gender | Female | 18 | 49% | 6 | 40% |
|  | Male | 18 | 49% | 9 | 60% |
|  | Prefer not to answer | 1 | 3% | 0 | 0% |
| Cause | Congenital | 23 | 62% | 12 | 80% |
|  | Acquired | 14 | 38% | 3 | 20% |
| Level | A | 2 | 5% | 2 | 13% |
|  | B | 26 | 70% | 10 | 67% |
|  | C | 9 | 24% | 3 | 20% |
| Prosthesis use | I use my prosthesis daily | 15 | 41% | 3 | 20% |
|  | I use my prosthesis at least once a week | 6 | 16% | 4 | 27% |
|  | I use my prosthesis less than once a week | 0 | 0% | 0 | 0% |
|  | I used a prosthesis in the past but not anymore | 6 | 16% | 4 | 27% |
|  | I never used a prosthesis | 10 | 27% | 4 | 27% |
| Duration of use | less than 1 year | 3 | 14% | 1 | 14% |
|  | 1 to 5 years | 5 | 24% | 2 | 29% |
|  | 6 to 10 years | 3 | 14% | 1 | 14% |
|  | more than 10 years | 10 | 48% | 2 | 29% |

#### B: Interview Participants

**Table 3:** Profiles of interview participants

| pseudo name | Level | Cause | Gender | Prostheses use | Years of use | Type of prostheses |
| --- | --- | --- | --- | --- | --- | --- |
| Ted | B | C | Male | I use my prosthesis at least once a week | 6 to 10 | BP, M |
| Paul | C | A | Male | I use my prosthesis at least once a week | >10 | M |
| Ross | B | C | Male | I never used a prosthesis |  |  |
| Kelsey | B | C | Female | I used a prosthesis in the past |  | M |
| Georgie | B | C | Male | I used a prosthesis in the past |  | BP |
| Jerome | B | C | Male | I use my prosthesis daily | >10 |  |
| Walter | C | A | Male | I never used a prosthesis |  |  |
| Tony | A | C | Male | I never used a prosthesis |  |  |
| Guy | B | C | Male | I used a prosthesis in the past |  | M |
| Cheryl | A | C | Female | I never used a prosthesis |  |  |
| Grace | B | C | Female | I used a prosthesis in the past |  |  |
| Margaret | B | A | Female | I use my prosthesis daily | <1 | BP, M |
| Laura | C | C | Female | I use my prosthesis at least once a week |  | C |
| Kevin | B | C | Male | I use my prosthesis at least once a week | 1 to 5 | M |
| Jessica | B | C | Female | I use my prosthesis daily | 1 to 5 | M |

#### C: Stakeholders’ Profiles

**Table 4:** Survey and Interviews Participant Demographics

|  |  |
| --- | --- |
| Prof Laurence Kenny | Rehabilitation research engineer, with over 20 years of experience in the field. |
| Kameron Maxwell | Dual qualified Prosthetist/Orthotist with experiencing working in a variety of clinical settings in the UK with both NHS and private healthcare services. He also has an MSc in Prosthetics & Orthotics. |
| Jim Ashworth-Beaumont | With prior training and experience as a mechatronic and control systems engineer, Jim has been a senior Orthotist/Prosthetist for over 20 years and been a senior clinician with a special interest in traumatic injury and neurorehabilitation at the NHS for 16 years. His postgraduate qualifications are in Health Studies and Neurorehabilitation. He has also experienced upper-limb amputation and was able to share his lived experience too. |

#### D: Survey Questions Flow Chart

Figure 8 shows the main sections of the survey. The section on sensory feedback starts by guiding the participants to think about vibrotactile feedback.

**Figure 8:**
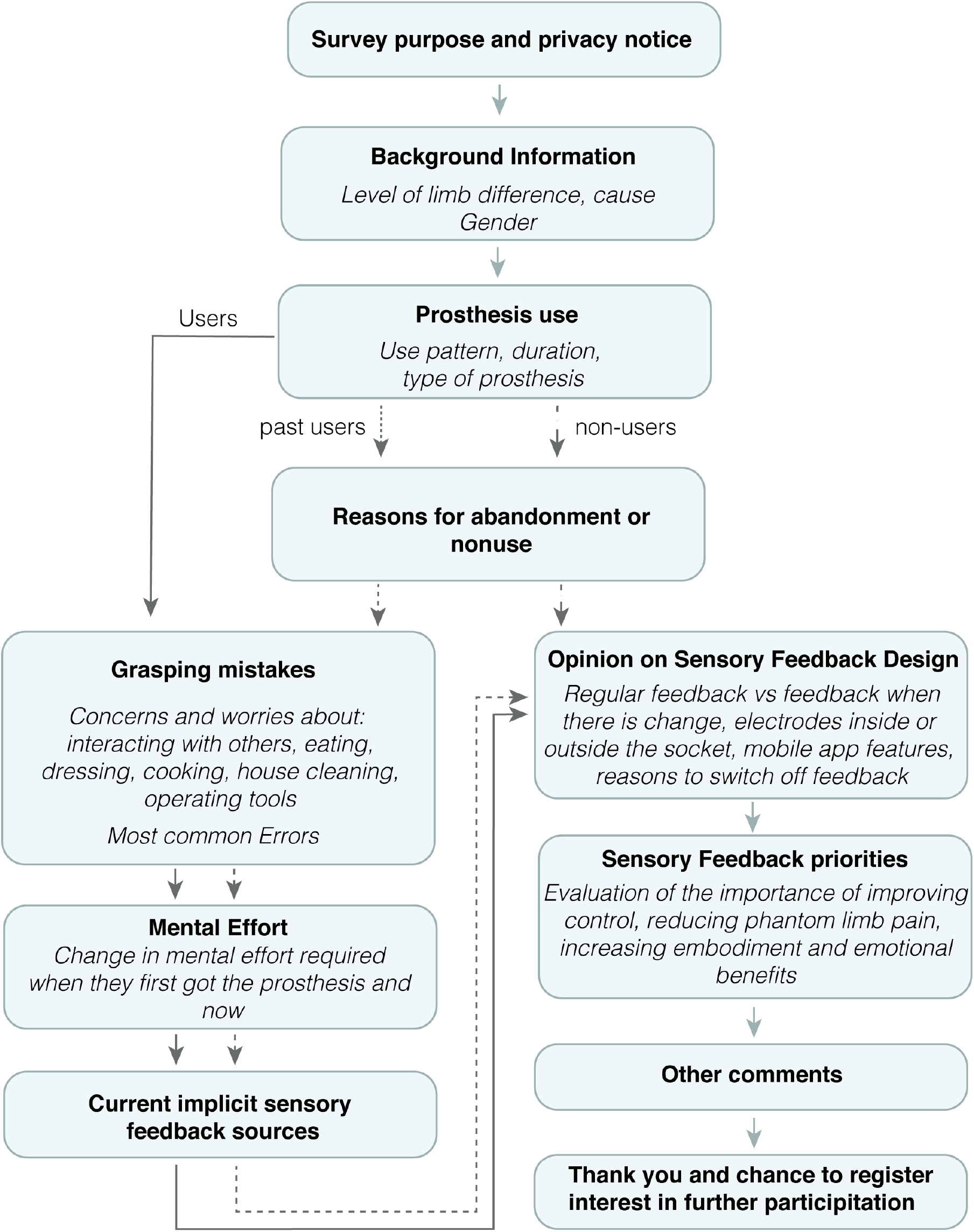
Flow chart showing the survey sections

## Acknowledgements

The authors would like to thank the participants and charities that helped with sharing the survey including: Blesma, The Douglas Bader Foundation, Limbless Association, I am possible foundation, Limbs 4 Life and E-Nable. The authors would also like to thank the stakeholders involved including Prof Laurence Kenney, Kameron Maxwell and Jim Ashworth-Beaumont. LJ would also like to thank Dr Alix Chadwell and Chantel Ostler for the thought provoking discussions and clinical insight.

## Abbreviations

ADL: Activities of Daily Living
PBA: Person Based Approach.

## Availability of data and materials

All of the available data is found in the additional material. The interview transcripts are treated as confidential and will not be shared.

## Ethics approval and consent to participate

This study has been approved by the University of Bath’s Psychology Research Ethics Committee (20-237).

## Consent for publication

Written consent was obtained from participants for the publication of the results.

## Authors’ contributions

Conceptualisation: LJ, BA, BWM, JR; Investigation: LJ, BA; Supervision: BWM, BA, DZ; Writing – original draft: LJ; Writing – review & editing: LJ, BA, BWM, JR, DZ

## Additional Files

### Additional file 1 — Additional Survey Information

This file includes the changes to the survey following the pilot studies as well as a summary of the additional results not reported in the paper. Those additional results cover worries and concerns about carrying out ADL with prostheses, opinion on frequency of feedback, electrode placement, mobile app and switching off feedback and overall ranking of sensory feedback priorities.

### Additional file 2 — Interview Schedule

This file includes the questions used to guide the interviews.

### Additional file 3 — Survey Questions

A copy of the online survey questions.

### Additional file 4 — Survey open text responses

A spreadsheet of the responses to open text questions.

### Additional file 5 — Interview codes

A list of the codes and themes generated with example quotes.

## Footnotes

1 Phantom limb pain describes neuropathic painful sensations experienced by individuals with acquired amputation. Those sensations are felt as though they are originating from the lost limb and can range from cramping to stabbing sensations. Individuals with congenital limb differences do not experience phantom limb pain, with the exception of some rare cases [10, 11].

2 The first pilot test resulted in substantial change to the survey so only the results from the second pilot study were included in the final results.

3 The profiles of the stakeholders consulted can be found in Appendix C.

4 widgets are a way for the user to use a mobile application through the home page without having to open the application itself with the aim of easier access to commonly used features.

## References

[1] R. F. Baumgartner, “Upper extremity amputation and prosthetics,” 2001.

[2] A. Saradjian, A. R. Thompson, and D. Datta, “The experience of men using an upper limb prosthesis following amputation: Positive coping and minimizing feeling different,” Disability and Rehabilitation, vol. 30, no. 11, pp. 871–883, 2008.

[3] I. C. Narang and V. S. Jape, “Retrospective study of 14,400 civilian disabled (new) treated over 25 years at an Artificial Limb Centre,” Prosthetics and Orthotics International, vol. 6, no. 1, pp. 10–16, 1982.

[4] D. J. Atkins, D. C. Y. Heard, and W. H. Donovan, “Epidemiologic overview of individuals with upper-limb loss and their reported research priorities,” JPO: Journal of Prosthetics and Orthotics, vol. 8, no. 1, pp. 2–11, 1996.

[5] S. Salminger, H. Stino, L. H. Pichler, C. Gstoettner, A. Sturma, J. A. Mayer, M. Szivak, and O. C. Aszmann, “Current rates of prosthetic usage in upper-limb amputees – have innovations had an impact on device acceptance?,” https://doi.org/10.1080/09638288.2020.1866684, 2020.

[6] K. Ostlie, R. J. Franklin, O. H. Skjeldal, A. Skrondal, and P. Magnus, “Musculoskeletal pain and overuse syndromes in adult acquired major upper-limb amputees,” Archives of Physical Medicine and Rehabilitation, vol. 92, no. 12, 2011.

[7] L. E. Jones and J. H. Davidson, “Save that arm: A study of problems in the remaining arm of unilateral upper limb amputees,” Prosthetics and Orthotics International, vol. 23, no. 1, pp. 55–58, 1999.

[8] J. B. Webster, N. Webster, M. Borgia, and L. Resnik, “Frequency, severity, and implications of shoulder pain in people with major upper limb amputation who use prostheses: Results of a National Study,” PM&R, pp. 1–11, 8 2021.

[9] M. A. Hanley, D. M. Ehde, M. Jensen, J. Czerniecki, D. G. Smith, and L. R. Robinson, “Chronic pain associated with upper-limb loss,” American journal of physical medicine and rehabilitation, vol. 88, pp. 742–751, 8 2009.

[10] H. Flor, “Phantom-limb pain: characteristics, causes, and treatment,” The Lancet Neurology, vol. 1, pp. 182–189, 7 2002.

[11] K. Limakatso, G. J. Bedwell, V. J. Madden, and R. Parker, “The prevalence of phantom limb pain and associated risk factors in people with amputations: a systematic review protocol,” Systematic Reviews 2019 8:1, vol. 8, pp. 1–5, 1 2019.

[12] S. J. Bensmaia, D. J. Tyler, and S. Micera, “Restoration of sensory information via bionic hands,” Nature Biomedical Engineering, 2020.

[13] “Sensory Feedback for Bionic Hands Bionics For Everyone.”

[14] E. Biddiss and T. Chau, “Upper-limb prosthetics: Critical factors in device abandonment,” American Journal of Physical Medicine and Rehabilitation, vol. 86, no. 12, pp. 977–987, 2007.

[15] C. Pylatiuk, S. Schulz, and L. Döderlein, “Results of an internet survey of myoelectric prosthetic hand users,” Prosthetics and Orthotics International, vol. 31, pp. 362–370, 12 2007.

[16] B. Stephens-Fripp, M. Jean Walker, E. Goddard, and G. Alici, “A survey on what Australians with upper limb difference want in a prosthesis: justification for using soft robotics and additive manufacturing for customized prosthetic hands,” Disability and Rehabilitation: Assistive Technology, vol. 15, no. 3, pp. 342–349, 2020.

[17] S. Lewis, M. F. Russold, H. Dietl, and E. Kaniusas, “User demands for sensory feedback in upper extremity prostheses,” in MeMeA 2012 - 2012 IEEE Symposium on Medical Measurements and Applications, Proceedings, pp. 188–191, 2012.

[18] E. L. Graczyk, A. Gill, D. J. Tyler, and L. J. Resnik, “The benefits of sensation on the experience of a hand: A qualitative case series,” PLoS ONE, vol. 14, no. 1, pp. 1–29, 2019.

[19] M. Y. Feilzer, “Doing Mixed Methods Research Pragmatically: Implications for the Rediscovery of Pragmatism as a Research Paradigm:,” http://dx.doi.org/10.1177/1558689809349691, vol. 4, pp. 6–16, 10 2009.

[20] D. L. Morgan, “Integrating Qualitative and Quantitative Methods: A Pragmatic Approach,” Integrating Qualitative and Quantitative Methods: A Pragmatic Approach, 12 2017.

[21] L. Yardley, B. Ainsworth, E. Arden-Close, and I. Muller, “The person-based approach to enhancing the acceptability and feasibility of interventions,” 2015.

[22] C. L. McDonald, C. L. Bennett, D. K. Rosner, and K. M. Steele, “Perceptions of ability among adults with upper limb absence: impacts of learning, identity, and community,” Disability and Rehabilitation, 2019.

[23] C. Lewis and J. Rieman, Task-centered User Interface Design: A Practical Introduction. University of Colorado, Boulder, Department of Computer Science, 1993.

[24] A. P. Field, Discovering statistics using IBM SPSS statistics : and sex and drugs and rock’n’roll. London: SAGE, 4th ed. ed., 2013.

[25] V. Braun and V. Clarke, “Using thematic analysis in psychology,” Qualitative Research in Psychology, vol. 3, no. 2, pp. 77–101, 2006.

[26] V. Braun and V. Clarke, “Reflecting on reflexive thematic analysis,” https://doi.org/10.1080/2159676X.2019.1628806, vol. 11, pp. 589–597, 8 2019.

[27] V. Braun and V. Clarke, “One size fits all? What counts as quality practice in (reflexive) thematic analysis?,” https://doi.org/10.1080/14780887.2020.1769238, vol. 18, no. 3, pp. 328–352, 2020.

[28] H. Torrance, “Triangulation, Respondent Validation, and Democratic Participation in Mixed Methods Research:,” http://dx.doi.org/10.1177/1558689812437185, vol. 6, pp. 111–123, 2 2012.

[29] A. Althubaiti, “Information bias in health research: definition, pitfalls, and adjustment methods,” Journal of Multidisciplinary Healthcare, vol. 9, p. 211, 5 2016.

[30] J. Zbinden, E. Lendaro, and M. Ortiz-Catalan, “Prosthetic embodiment: Review and perspective on definitions, measures, and experimental paradigms.” (preprint), 4 2021.

[31] J. Zbinden, E. Lendaro, and M. Ortiz-Catalan, “A multi-dimensional framework for prosthetic embodiment: Review and perspective for translational research.” (preprint), 2 2022.

[32] K. Østlie, I. M. Lesjø, R. J. Franklin, B. Garfelt, O. H. Skjeldal, and P. Magnus, “Prosthesis use in adult acquired major upper-limb amputees: patterns of wear, prosthetic skills and the actual use of prostheses in activities of daily life,” Disability and Rehabilitation: Assistive Technology, vol. 7, pp. 479–493, 11 2012.

[33] N. Kerver, S. v. Twillert, B. Maas, and C. K. v. d. Sluis, “User-relevant factors determining prosthesis choice in persons with major unilateral upper limb defects: A meta-synthesis of qualitative literature and focus group results,” PLOS ONE, vol. 15, p. e0234342. 6 2020.

[34] J. Y. Zheng, C. Kalpakjian, M. Larrága-Martínez, C. A. Chestek, and D. H. Gates, “Priorities for the design and control of upper limb prostheses: A focus group study,” Disability and Health Journal, vol. 12, pp. 706–711, 10 2019.

[35] L. C. Smail, C. Neal, C. Wilkins, and T. L. Packham, “Comfort and function remain key factors in upper limb prosthetic abandonment: findings of a scoping review,” Disability and Rehabilitation: Assistive Technology, vol. 0, no. 0, pp. 1–10, 2020.

[36] S. M. Engdahl, B. P. Christie, B. Kelly, A. Davis, C. A. Chestek, and D. H. Gates, “Surveying the interest of individuals with upper limb loss in novel prosthetic control techniques,” Journal of NeuroEngineering and Rehabilitation, vol. 12, no. 1, pp. 1–11, 2015.

[37] C. Dietrich, K. Walter-Walsh, S. Preißler, G. O. Hofmann, O. W. Witte, W. H. Miltner, and T. Weiss, “Sensory feedback prosthesis reduces phantom limb pain: Proof of a principle,” Neuroscience Letters, vol. 507, no. 2, pp. 97–100, 2012.

[38] S. Preißler, D. Thielemann, C. Dietrich, G. O. Hofmann, W. H. Miltner, and T. Weiss, “Preliminary evidence for training-induced changes of morphology and phantom limb pain,” Frontiers in Human Neuroscience, vol. 11, no. June, pp. 1–12, 2017.

[39] P. M. Rossini, S. Micera, A. Benvenuto, J. Carpaneto, G. Cavallo, L. Citi, C. Cipriani, L. Denaro, V. Denaro, G. Di Pino, F. Ferreri, E. Guglielmelli, K. P. Hoffmann, S. Raspopovic, J. Rigosa, L. Rossini, M. Tombini, and P. Dario, “Double nerve intraneural interface implant on a human amputee for robotic hand control,” Clinical Neurophysiology, vol. 121, no. 5, pp. 777–783, 2010.

[40] F. M. Petrini, G. Valle, I. Strauss, G. Granata, R. Di Iorio, E. D’Anna, P. Čvančara, M. Mueller, J. Carpaneto, F. Clemente, M. Controzzi, L. Bisoni, C. Carboni, M. Barbaro, F. Iodice, D. Andreu, A. Hiairrassary, J. L. Divoux, C. Cipriani, D. Guiraud, L. Raffo, E. Fernandez, T. Stieglitz, S. Raspopovic, P. M. Rossini, and S. Micera, “Six-Month Assessment of a Hand Prosthesis with Intraneural Tactile Feedback,” Annals of Neurology, vol. 85, no. 1, pp. 137–154, 2019.

[41] D. W. Tan, M. A. Schiefer, M. W. Keith, J. R. Anderson, J. Tyler, and D. J. Tyler, “A neural interface provides long-term stable natural touch perception,” Science Translational Medicine, vol. 6, no. 257, 2014.

[42] A. Chadwell, L. Kenney, S. Thies, J. Head, A. Galpin, and R. Baker, “Addressing unpredictability may be the key to improving performance with current clinically prescribed myoelectric prostheses,” Scientific Reports 2021 11:1, vol. 11, pp. 1–15, 2 2021.

[43] C. Widehammar, I. Pettersson, G. Janeslätt, and L. Hermansson, “The influence of environment: Experiences of users of myoelectric arm prosthesis—a qualitative study,” Prosthetics and Orthotics International, vol. 42, pp. 28–36, 2 2018.

[44] K. Malterud, V. D. Siersma, and A. D. Guassora, “Sample Size in Qualitative Interview Studies: Guided by Information Power,” Qualitative Health Research, vol. 26, pp. 1753–1760, 11 2016.

