## Additional file 1: Additional Survey information for "Experience of Adults with Upper-limb Difference and their Views on Sensory Feedback for Prostheses: A Mixed Methods Study"

### Survey Additional File

#### Changes to Survey questions following the two piloting tests

**Table 1 Summary of changes made to the survey based on the piloting tests with two participants, P(i) and P(ii)**

| Section | Change | Rationale |
|---|---|---|
| Background information | "Bilateral" to "Both hands" | P(i) was not sure what bilateral meant |
| | Added a note to the prostheses use section highlighting that the answer should reflect normal use of the prostheses before COVID | P(i)'s prosthesis use changed during COVID19 due to the difficulty of cleaning the prosthesis properly when going out. |
| | Cause of limb loss "Congenital" was changed to "Congenital (from birth) | P(ii) pointed out that some may not know what congenital means. |
| Grasping mistakes | The first question was on what the most common errors are made when using a prosthesis. This was moved further to follow the questions on specific tasks. | P(i) was confused about what "errors" meant. |
| | The question on specific grasping mistakes was a multiple-choice question. This was changed to be an open text question where the participants can mention any worries they have about using the prosthesis for the tasks. | P(i) found this to be a confusing question and wasn't sure what the purpose was. Changing it to open text enables participants to answer it more easily without trying to understand the different options. |
| Opinion on sensory feedback design | Deleted the question "Could you please give an example of times when you do not think that you need to have sensory feedback and explain why?" and | P(i) pointed out that the question is too similar to the following one "The sensory feedback system will have an option to switch it off. When do you expect to want to switch it off and what would be the reason for that?" which caused confusion. |

#### Prosthesis Experience

The most common reasons for not using a prosthesis were comfort and not needing one. Lack of sensation followed as the third most common reason (see Figure 6 in main text). Participants were able to provide other reasons using an open text box. Most of the responses were reworded versions of the available options except one participant who was waiting to receive his prosthesis.

Prosthesis users reported that they often worry about safety when interacting with others (ex. holding babies and shaking hands) as well as when cooking (ex. fear of it being a fire hazard). Eating seemed to be one of the difficult tasks to do with a prosthesis due to the limitations of elbow and wrist movement as well as the difficulties of holding a knife securely when cutting food. When asked about the most common errors, 37.5% of the participants mentioned errors related to holding objects such as dropping them or holding them too tightly. Other errors included unexpected movements, cycling through grips and orientation errors. Some participants mentioned that they sometimes tend to use their prostheses in an unconventional manner. For example:

> *"When confronted with a new task I don't do it the "right" way (utilizing the prosthetic hand) well if I'm in a hurry. I push objects against my arm like I would with no prosthetic."*

(P32, Level 4 congenital limb difference, using a Myoelectric prosthesis for 5 years)

*"I almost never use the opening and closing function because I'll always over squeeze or drop things. I know I should practice more so as to not use it so passively but I'm [in my 30s] so I doubt I'll learn new tricks at this point but it's possible"*

(P33, Level 5 congenital limb difference, using a Myoelectric prosthesis for 31 years)

**Table 2 Worries and concerns about ADL**

| | |
|---|---|
| Interacting with kids/ babies | •Safety (hard, difficult to pick up effectively)<br>•Looks scary |
| Interacting with others | •Safety (not being able to release)<br>•Prejudice<br>•Making others uncomfortable |
| Eating | •Cutting food (knife drops out)<br>•Difficult without elbow/wrist movement<br>•Dropping objects |
| Dressing | •Fitting through sleeves |
| Cooking | •Safety (fire hazard)<br>•Water/heat damage |
| House cleaning | •None |
| Operating tools | •Holding screws/ nails |
| Common errors | •Dropping objects/ losing grip<br>•Holding too tight<br>•Unexpected movements (ex. due to electrode lift-off)<br>•Not always used the "right way" especially with new tasks |

#### Opinion on Sensory Feedback Design

When given the choice between regular feedback and feedback when there is change, 62.2% chose the latter (Figure 1) with the justification that constant feedback can be overstimulating or turn into white noise. This might result in feeling like they are losing control or cause confusion. A few also mentioned the effect this might have on battery life. Those who wanted regular feedback explained that they want to be able to determine the grip strength and know that the feedback is working.

**Table 3 Justification for type of feedback chosen**

| When there is change | Regular Feedback |
|---|---|
| - Constant feedback results in it becoming white noise<br>- Constant feedback is overstimulating<br>- More in control<br>- Reduce confusion<br>- Battery life<br>- Could cause me to drop what I am holding | - To determine grip strength<br>- To know it is working correctly<br>- Can see changes but not intensity<br>- Consistency |

The second question was on the location of the electrodes used to provide the feedback. 67.5% preferred the electrodes to be inside the socket as it is tidier and will not require the user to position them. It was also pointed out that it might be a more intuitive location for the electrodes to be. Those who selected using a cuff instead did so as they thought it provides greater flexibility or were aware of the limited space available in the socket.

The introduction of a mobile app to modify the settings received mostly positive feedback with a wide range of features suggested such as feedback training, measurement of prosthesis usage, visualisation of grip strength and intensity adjustment

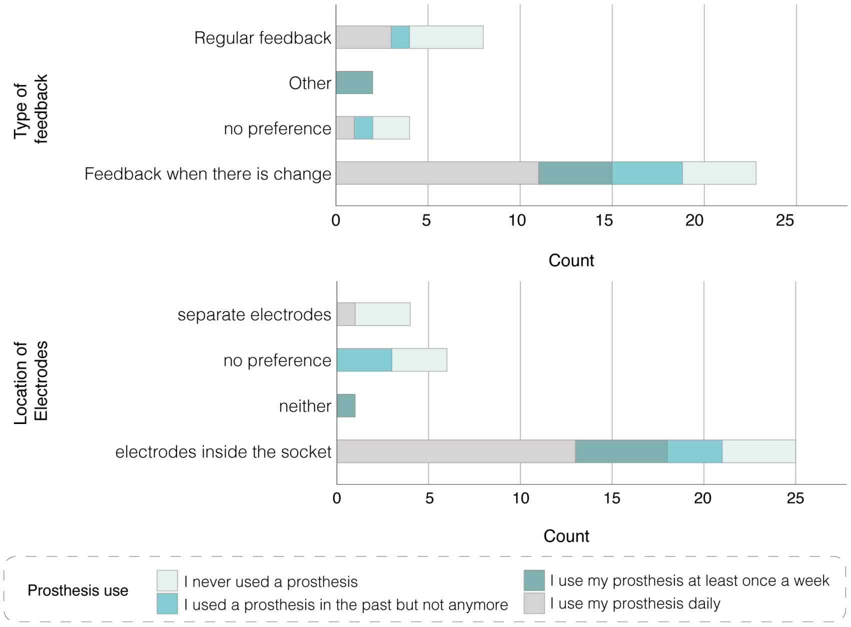


**Figure 1** The participant preferences for the type of feedback and location of electrodes split based on prosthesis use.

**Table 4** Justification for location of electrodes

| Inside the socked | As a cuff |
|---|---|
| - More tidy<br>- Interference with roll on sleeve<br>- Difficult for bilateral amputees<br>- Will trust the positioning more if it was inside the socket<br>- might be more intuitive<br>- avoid additional width/ difficulty with clothing | - More flexible fit<br>- Small area (of socket)<br>- Interference with socket pressure |

(see Table 5). The concerns were mostly related to connectivity issues with the importance of alternative options being highlighted. The need for the app to be simple and easily compatible was also mentioned.

**Table 5** Open text responses about the mobile app

| Concerns | Desired features |
|---|---|
| - Time needed<br>- Alternative option in the case of connection issues/ losing phone<br>- Difficult to use an app with one hand<br>- updates and compatibility | - Limited options for simplicity<br>- Lots of sequence patterns<br>- Intensity adjustment<br>- voice control<br>- battery life<br>- Arm information<br>- Feedback training<br>- Test mode<br>- measuring usage of prosthesis<br>- visualisation of grip strength<br>- sliding scales |

The reasons for switching off sensory feedback reinforced the thoughts presented in the previous questions. Participants seem to be worried about feedback becoming intrusive and specified wanting to switch feedback off when holding an object for a long time or when trying to rest. Passive prosthesis use and use with simple tasks where the user feels in control were other examples. Switching feedback off to conserve battery power was also mentioned.

**Table 6** Open text responses about switching the feedback off

| **Reasons for switching feedback off** |
| --- |
| - During a tight grip for a long time |
| - When trying to rest (reading, watching TV) |
| - In a quiet place (if there is sound) |
| - Light duty chores (when not worried about grip strength) |
| - To conserve power |
| - When feeling in control |
| - Passive use |

##### Opinion on Sensory Feedback Priorities

Participants were asked to evaluate how important each of the potential benefits of sensory feedback is *to them*. The benefits included improving control, reducing phantom limb pain, increasing embodiment and emotional benefits. The scoring was on a Likert scale between 1 (not important) and 4 (very important). Participants with acquired limb differences ranked reducing phantom limb pain as the highest priority, followed by improving control. Most participant with congenital limb differences do not experience phantom limb pain and, therefore, did not consider it an important feature and ranked their top priority as improving control. It is interesting to note that improving control was ranked significantly lower by participants with acquired limb differences. This may be due to the importance of reducing phantom limb pain being reflected in how they ranked improving control (i.e., it being less important than reducing phantom limb pain). Alternatively, this could reflect that the existing level of control is considered effective for their needs. Increasing embodiment and emotional benefits received lower importance with emotional benefits being the least important (see Figure 5 in the main text).

The justifications for the importance of improving control reflected the most common errors reported in the previous sections. Participants wanted to be able to have a better understanding of what the prosthesis is doing to be able to trust it. This trust is thought to help with not worrying about hurting others unintentionally or dropping/ crushing objects while holding them. P16 linked the significance of improved prosthesis control to better control over her life *"That way we can feel we have control over our lives"*. The importance of embodiment varied between individuals with those ranking it highly doing so because they thought it would result in more intuitive use and increased confidence. Those who ranked it lower, on the other hand, did so because they viewed prostheses as tools. Similar comments were made on the importance of emotional benefit. contrary to expectations, there was no significant difference between how participants with acquired and congenital limb difference ranked the emotional benefit of sensory feedback. Those who ranked it highly did so because they thought it would encourage them to wear the prosthesis more. Interestingly, participants with congenital limb differences were curious about how that would feel, particularly because they never "felt" through their missing limb. Similar to the embodiment justifications, those who viewed the prosthesis as a tool were not interested in the emotional benefits, with P2 mentioning that *"[his] emotions don't work that way."*.

P28 commented that trying sensory feedback had had a negative effect on his phantom limb pain.

> *"I had that for a study once and it really exacerbates my phantom pains beyond any measure, so I dropped out."*

**Table 7** Open text responses about priorities in relation to sensory feedback

| Priority | Main points | Quotes |
|---|---|---|
| **Control** | •Don't want to hurt people by accident<br>•Trusting the control of the prosthesis<br>•Helps understand what the prosthesis is doing better<br>•Don't want to drop/ crush objects | *"whole point of having one"* |
| **Phantom limb pain** | • very tiring and painful<br>•Because it can be emotionally distressing to feel these types of pain | |
| **Embodiment** | • Intuitive use<br>•Confidence<br>•Those who said its not important did that because they view it as a tool | *"I want it to be an [extension] of me verses a movie arm" (never used one)* |
| **Emotional benefit** | • Makes it feel less like a tool (positive)<br>•Might encourage users to wear the prosthesis more<br>•Congenital amputees split between not caring or being curious because they never felt touch through missing hand. | *"Not sure whether this would be the case. Could have opposite effect - remind you that you've lost a limb."* |
