## Additional file 2:Interview schedule for "Experience of Adults with Upper-limb Difference and their Views on Sensory Feedback for Prostheses: A Mixed Methods Study"

### Interview Appendix

#### Interview questions

The interviews started by introductions and a brief chat to establish rapport. Once the participant is ready, the recording is started and the questions below are used to guide the conversation, when suitable.

- Tell me about your experience with limb difference. Do you wear a prosthesis?
  - How did it feel when you first got it?
  - How did this feeling change with time?
  - Did social interaction with friends, family and health professionals play a role in your rehabilitation process? [ask them to elaborate]
- In your day-to-day life, what determines whether or not you will wear the prostheses?
  - Did those factors change since you started wearing the prostheses? - Why?
  - What do you think the benefits of wearing it are?
- The last section of the survey you filled was about sensory feedback. Have you thought about sensory feedback before?
- I would just like to expand on the questions we asked in the survey
- If you were given the option to add sensory feedback to your prostheses, what would be some of the things you would think about?
  - How would you use it?
  - What would be the first thing you would want to try with sensory feedback
  - How do you expect it to work
  - What would you say are the most important features
  - What would you be worried about?
  - How do you feel about having to learn how to use the sensory feedback system? (often followed by explaining that non-invasive approaches require learning what certain patterns mean and the use of a mobile app to adjust settings)
  - When would you have it on or off?
  - Is there anything else you would want to add?
