## Additional file 3: Survey questions for "Experience of Adults with Upper-limb Difference and their Views on Sensory Feedback for Prostheses: A Mixed Methods Study"

### Control of Prostheses

#### Survey purpose and privacy notice

Upper-limb prostheses can be difficult to use due to many reasons. Researchers at the University of Bath are working on developing a system to improve the control of upper-limb prostheses. Including the users in the design process enables for more effective solutions to be implemented. Therefore, the aim of this study is to understand the users' perspective on some aspects of prostheses use and get their opinion on proposed design features.

Upon completion of this survey, you will be invited to register your interest in further involvement in the research **through a link**. This includes participation in video interviews, lab testing of prototype devices and home testing of prototype devices. You can choose whichever of the activities you are interested in or none.

If you have any questions or concerns, please feel free to contact the researchers working on this project:

Researcher: Leen Jabban

Supervisors: Dr Benjamin Metcalfe, Dr Ben Ainsworth and Dr Dingguo Zhang

Psychology Research Ethics Committee contact:

**Please read the following statements carefully and choose if you agree with them or not.**

| |
|---|
| I am over 18 years old. |
| I am an upper-limb amputee. |
| I have received enough information about the project to make a decision about my participation. |
| I understand the nature and purpose of the procedures involved in this project. These have been communicated to me on the information sheet accompanying this form. |

| |
|---|
| I understand that the University of Bath may use the data collected for this study in future research project(s) but that the conditions on this form under which I have provided the data will still apply. |
| I understand the data I provide will be treated as confidential, and that no identifying information will be stored in relation to the survey responses. |
| I agree to the University of Bath keeping and processing the data that I provide during the course of this study and my consent is conditional upon the University complying with its duties and obligations under the Data Protection Act |
| I hereby fully and freely consent to my participation in this project. |

I agree with **all** of the above statements

- Yes
- No

### Background information

Which of the levels defined in the image below matches your level of limb loss?
✱ *Required*

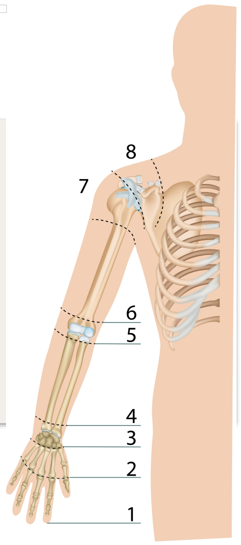

- 1
- 2
- 3
- 4
- 5
- 6
- 7
- 8
- 9

What was the cause of your limb loss? ✱ *Required*

- Trauma
- Congenital (from birth)
- Other

If you selected Other, please specify:

For which hand has your limb loss occurred?

- Dominant hand
- Non-dominant hand
- Both

Gender ✱ *Required*

- Male
- Female
- Other
- Prefer not to answer

### Prostheses use

Which statement matches how you would describe your use of a prosthesis?
Note: If COVID19 has influenced your prosthesis use please answer this question to reflect how you used it **before** COVID19. ✱ *Required*

- I use my prosthesis daily
- I use my prosthesis at least once a week
- I use my prosthesis less than once a week
- I used a prosthesis in the past but not anymore
- I never used a prosthesis

How long have you been using a prosthesis for?

Which type of prostheses do you use?

Why did you choose not to use a prosthesis? (you may select multiple answers)

- It is not comfortable
- It is hard to use
- It is expensive to buy
- It is expensive to maintain
- I cannot feel what I am doing through it
- I do not need one
- Other

If you selected Other, please specify:

#### Grasping Mistakes

What are some concerns or worries you have when you attempt to carry out the following tasks with a prosthesis? If you do not use a prosthesis to carry out the task please state that.

| | ✱ *Required* |
|---|---|
| Interacting with kids/ babies (e.g. holding them) | |
| Interacting with others (e.g. shaking hands) | |
| Eating | |
| Dressing | |
| Cooking | |
| House cleaning | |
| Operating tools | |

What would you say are the most common errors you make when using a prosthesis?

How hard do you find using the prosthesis hand in terms of thinking, planning your actions and remembering commands? Please select a value from 0 to 5 with 0 being low effort and 5 being high effort. Examples: 0: I can use the prosthesis and maintain a conversation without interruption 5: I cannot do anything else while trying to use the prosthesis ✱ *Required*

Please don't select more than 1 answer(s) per row.

Please select at least 2 answer(s).

| | 0 (low) | 1 | 2 | 3 | 4 | 5 (high) |
|---|---|---|---|---|---|---|
| When you first got it | ☐ | ☐ | ☐ | ☐ | ☐ | ☐ |
| now | ☐ | ☐ | ☐ | ☐ | ☐ | ☐ |

How does that affect your use of the prosthesis?

### Sensory feedback

How do you know that the prosthesis is applying the force you want it to? **Please select all that apply.** ✱ *Required*

Please select at least 1 answer(s).

- [ ] I look at the arm
- [ ] I listen to the sound of the motors
- [ ] I feel the vibration through the socket
- [ ] I don't know
- [ ] Other

If you selected Other, please specify:

If you selected more than one way, which one would you say is the one you rely on most? why?

### Opinion

We are working on developing a sensory feedback system for upper-limb prostheses. The system will rely on placing electrodes on the residual limb. Those electrodes will provide you with a sensation that you can use as an indication of how the prosthesis is engaging with the object held (for example, I how hard you are holding an object).

The aim of the following questions is to get your opinion on the system. The comments you provide will help guide the design process.

The feeling you will get through the sensory feedback system will be similar to the feeling of your phone vibrating when you get a notification.   Imagine that you are trying to hold an object, a cup for example. Would you prefer to get the vibration feeling when there is a change (for example, the cup is about to fall/slip) or would you prefer to get regular vibration (for example, vibration every few seconds that changes in intensity based on how hard you are holding the cup)? ✱ *Required*

- ○ Feedback when there is change
- ○ Regular feedback
- ○ no preference
- ○ Other

If you selected Other, please specify:

________________

Why?

________________

What are other changes in how the prosthesis is engaging with the object held (other than slip) when getting this sensory feedback could be helpful?

________________

In order to deliver the feeling, electrodes will have to be placed on your skin. You will feel the vibration through the electrodes. Would you prefer to have the electrodes integrated into the socket or separate (a cuff, for example, with wireless connection)?

- electrodes inside the socket
- separate electrodes
- no preference
- neither

Could you please explain why? What are some concerns you have about the electrode location?

The system will enable you to modify the sensation settings using a mobile app. What are some concerns that you have about the way you can adjust the sensation settings?

What are some features you hope to see in the app?

The sensory feedback system will have an option to switch it off. When do you expect to want to switch it off and what would be the reason for that? ✱ *Required*

The following benefits have been associated with sensory feedback. Could you please rate them based on how important they are **to you**? Your opinion will enable us to know what to focus on as part of our research.

| | ✱ *Required* | | | | |
|---|---|---|---|---|---|
| | not important | neutral | important | very important | Why? ✱ *Required* |

| | | | | |
|---|---|---|---|---|
| Makes it easier to control the prostheses | ◯ | ◯ | ◯ | ◯ |
| Reduces phantom limb pain | ◯ | ◯ | ◯ | ◯ |
| Makes the prosthesis feel more like a part of the body rather than a tool | ◯ | ◯ | ◯ | ◯ |
| It is emotionally pleasing to feel touch through the prosthesis | ◯ | ◯ | ◯ | ◯ |

Is there anything we haven't mentioned?

Do you have any extra comments or concerns?

We are looking for **participants to get involved in** other parts of the research such as **interviews** and **testing of prototypes** in the lab and/or **at home**.

You can register your interest using the link that will be shown once you submit the survey.

### Thank you

Thank you for participating in our survey on the control of upper-limb prostheses. Your input will guide our design of a sensory feedback system.

We are looking for participants to get involved in other parts of the research such as interviews and testing of prototypes in the lab and/or at home. If you would like to be considered, please fill in your contact details using the link below:

https://bathreg.onlinesurveys.ac.uk/research-participation

Once again, we sincerely thank you and appreciate your time and dedication to participate in our online survey.

If you have any questions on this study, please contact us. Our contact details can be found below:

Researcher: Leen Jabban

Supervisors: Dr Benjamin Metcalfe, Dr Ben Ainsworth and Dr Dingguo Zhang

Psychology Research Ethics Committee contact:

**Thank you again for your participation.**
